# Modelling the impact of reopening schools in the UK in early 2021 in the presence of the alpha variant and with roll-out of vaccination against SARS-CoV-2

**DOI:** 10.1101/2021.02.07.21251287

**Authors:** J. Panovska-Griffiths, R.M. Stuart, C.C. Kerr, K. Rosenfield, D. Mistry, W. Waites, D.J. Klein, C. Bonell, R.M. Viner

## Abstract

**Background:** Following the resurgence of the COVID-19 epidemic in the UK in late 2020 and the emergence of the alpha (also known as B117) variant of the SARS-CoV-2 virus, a third national lockdown was imposed from January 4, 2021. Following the decline of COVID-19 cases over the remainder of January 2021, the question of when and how to reopen schools became an increasingly pressing one in early 2021. This study models the impact of a partial national lockdown with social distancing measures enacted in communities and workplaces under different strategies of reopening schools from March 8, 2021 and compares it to the impact of continual full national lockdown remaining until April 19, 2021.

**Methods:** We used our previously published agent-based model, Covasim, to model the emergence of the alpha variant over September 1, 2020 to January 31, 2021 in presence of Test, Trace and Isolate (TTI) strategies. We extended the model to incorporate the impacts of the roll-out of a two-dose vaccine against COVID-19, with 200,000 daily vaccine doses prioritised by age starting with people 75 years or older, assuming vaccination offers a 95% reduction in disease acquisition risk and a 30% reduction in transmission risk. We used the model, calibrated until January 25, 2021, to simulate the impact of a full national lockdown (FNL) with schools closed until April 19, 2021 versus four different partial national lockdown (PNL) scenarios with different elements of schooling open: 1) staggered PNL with primary schools and exam-entry years (years 11 and 13) returning on March 8, 2021 and the rest of the schools years on March 15, 2020; 2) full-return PNL with both primary and secondary schools returning on March 8, 2021; 3) primary-only PNL with primary schools and exam critical years (years 11 and 13) going back only on March 8, 2021 with the rest of the secondary schools back on April 19, 2021 and 4) part-rota PNL with both primary and secondary schools returning on March 8, 2021 with primary schools remaining open continuously but secondary schools on a two-weekly rota-system with years alternating between a fortnight of face-to-face and remote learning until April 19, 2021. Across all scenarios, we projected the number of new daily cases, cumulative deaths and effective reproduction number R until April 30, 2021.

**Results:** Our calibration across different scenarios is consistent with alpha variant being around 60% more transmissible than the wild type. We find that strict social distancing measures, i.e. national lockdowns, were essential in containing the spread of the virus and controlling hospitalisations and deaths during January and February 2021. We estimated that a national lockdown over January and February 2021 would reduce the number of cases by early March to levels similar to those seen in October 2020, with R also falling and remaining below 1 over this period. We estimated that infections would start to increase when schools reopened, but found that if other parts of society remain closed, this resurgence would not be sufficient to bring R above 1. Reopening primary schools and exam critical years only or having primary schools open continuously with secondary schools on rotas was estimated to lead to lower increases in cases and R than if all schools opened. Without an increase in vaccination above the levels seen in January and February, we estimate that R could have increased above 1 following the reopening of society, simulated here from April 19, 2021.

**Findings:** Our findings suggest that stringent measures were integral in mitigating the increase in cases and bringing R below 1 over January and February 2021. We found that it was plausible that a PNL with schools partially open from March 8, 2021 and the rest of the society remaining closed until April 19, 2021 would keep R below 1, with some increase evident in infections compared to continual FNL until April 19, 2021. Reopening society in mid-April, without an increase in vaccination levels, could push R above 1 and induce a surge in infections, but the effect of vaccination may be able to control this in future depending on the transmission blocking properties of the vaccines.

## Introduction

At the end of January 2021, the UK remained severely affected by the global COVID-19 pandemic, with over 3.8 million confirmed cases and 100 thousand deaths having been recorded since the virus was first detected in the UK more than a year prior [1].

The increase in the number of COVID-19 cases over the period from November 2020 until January 2021 has been attributed to the emergence of new variants of SARS-CoV-2 [2]. One particular variant, called alpha variant but also known as the B117 variant, has been of particular interest [2,3], with studies suggesting it is notably more transmissible that other strains of SARS-CoV-2 [4,5]. WHO reports from early December 2020 suggest that in the period from October 5 to December 13, 2020, over 50% of COVID-19 cases in South-East England were identified as involving the alpha variant [2], with retrospective analysis suggesting that the new variant had been present in South-East England from September 20, 2020 [2].

As a result of the increase in COVID-19 cases across the UK over October and November 2020, a national lockdown was imposed between November 5, 2020 and December 3, 2020 (Figure 1) [6]. Following the release of this lockdown, COVID-19 cases, hospitalisations and deaths increased sharply during December 2020, prompting the UK Government to impose even stricter national lockdown conditions from January 5, 2021 [7] - the third such lockdown that the UK had implemented. During this third national lockdown, unlike during the November 2020 lockdown, schools remained closed apart from attendance by vulnerable children and the children of key workers.

**Figure 1:**
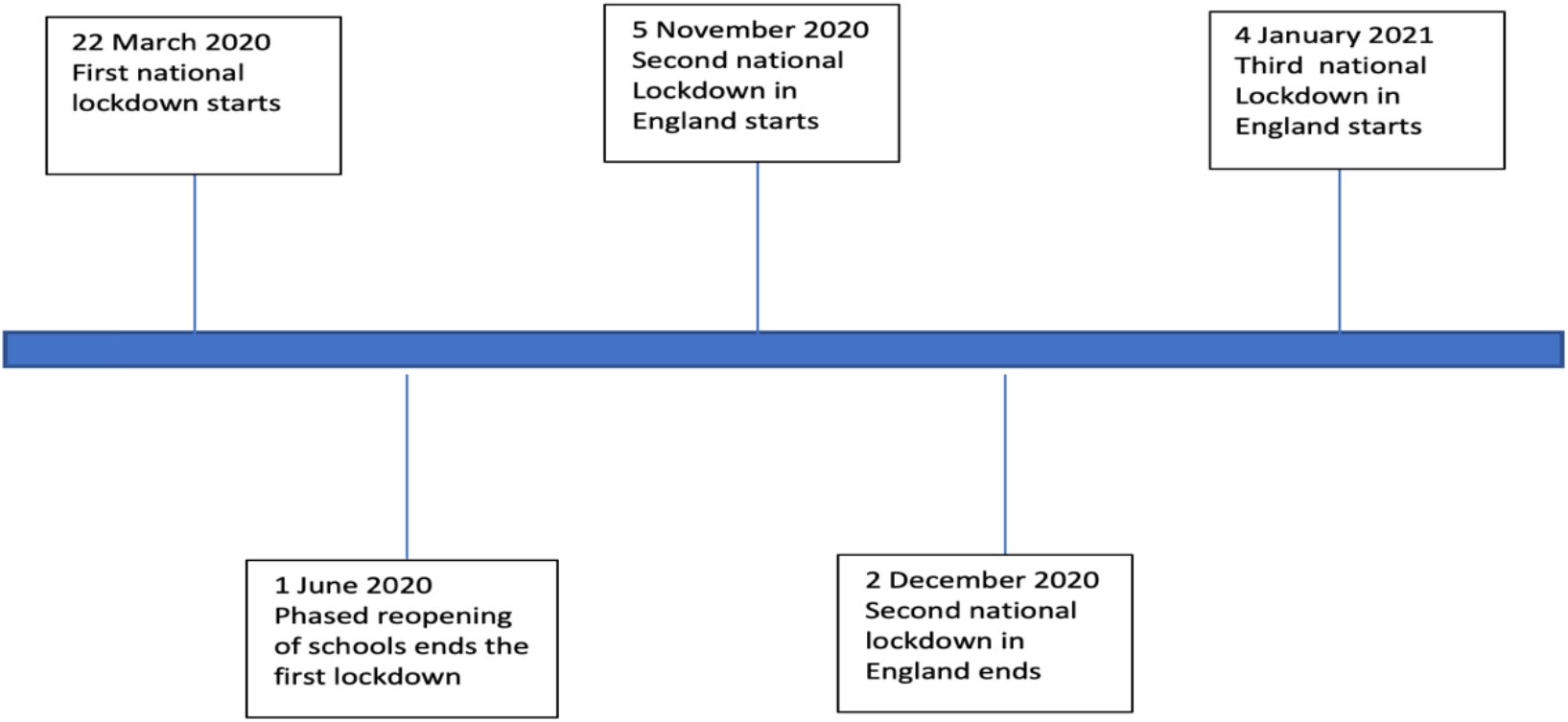
Timeline of the modelled COVID-19 associated lockdowns in the UK between March 2020 and January 2021.

In late December 2020, a national program of vaccination against COVID-19 began across the UK, with initial phases targeting elderly people and high-priority groups such as health-care workers [8]. In its initial stages, this vaccination program included two vaccines, the Pfizer/BioNTech and the Oxford/AstraZeneca vaccines, both two-dose regimes that were approved in the UK in late 2020. The current understanding is that these vaccines offer protection against severe disease with efficacy between 70-95% [9]. The expectation is that vaccination would reduce the rate of hospitalisations and deaths in the vaccinated cohort. There are still a number of open questions regarding the efficacy of the vaccines (including how much vaccination also reduces onward transmission [10,11], whether the vaccines achieve sterilising immunity and how long the effect of the vaccine lasts), many of which will only be answerable after sufficient time has elapsed to see these effects in vaccinated cohorts. However, in the absence of definitive data, mathematical modelling can help understand the possible impact of vaccines with various properties on the epidemic trajectory.

Throughout January 2021, the UK Government indicated that the third UK national lockdown would remain in place while the roll-out of the vaccination campaign continued and until a clear indication was received that the COVID-19 surge was under control [12]. At the same time, the UK Government has also emphasised its position that schools should be the first areas of society to reopen [13]. Apart from the considerable damage to children and young people’s learning [14], school closures can induce developmental, physical and mental health issues arising from lost education [15] as well as social isolation and reduced social support [16] and increased violence [17], as some examples of spillover effects. All of these factors are likely to present greater challenges in poorer families, consequently exacerbating inequalities. It is therefore critical to understand how various scenarios of partial or full reopening of schools in the presence of continued broader social lockdown might impact upon infections, deaths and reproduction number R in the presence of the alpha variant.

To answer this question, we used mathematical modelling to simulate the impact of a full national lockdown (FNL) in England from January 4, 2021 compared to partial national lockdowns (PNL) in which some elements of in-person schooling remained open. We assessed future epidemic trajectories until the end of April 2021 in five different scenarios: 1) **FNL** between January 4, 2021 and April 19, 2021 with all schools closed for in-person teaching and all but vulnerable students undertaking online learning, together with comprehensive restrictive social distancing measures across society; 2) **Staggered PNL** between January 4 and March 8, 2021 with schools open for face-to-face learning from March 8, 2021 for primary schools and for years 11 and 13 of secondary schools (being exam-critical years) and the rest of secondary school years from March 15, with online learning provided for secondary students between January 4 and the start of face-to-face learning; 3) **Full-return PNL** with both primary and secondary schools returning on March 8, 2021; and 4) **Primary-only PNL** with primary schools and exam critical year (years 11 and 13) only opened on March 8, 2021 with the rest of the secondary schools back on April 19, 2021 and 5) **Part-Rota PNL** with both primary and secondary schools returning on March 8, 2021 with primary students remaining in school continuously while the secondary students have two-weekly rota-system with face-to-face teaching provided for two weeks off until April 19, 2021.

All four PNL strategies within this analysis are accompanied by comprehensive social distancing measures across society and ongoing Test, Trace and Isolate interventions. All strategies also assume that there will be a two-week holiday for all schools from April 1-April 18 and that schools are ‘dismissed’ rather than closed i.e. that those not in classrooms are provided with access to online learning and that face-to-face teaching is available for vulnerable children and children of key workers. In January 2021, estimates were that around 1 in 5 primary school students were attending schools while only 5% of secondary school students were attending [18]. For each scenario, we projected the number of new daily cases, cumulative deaths and effective reproduction number (R).

## Methods

### Transmission model

We used Covasim, a stochastic individual-based model of SARS-CoV-2 transmission across a population [21]. We have previously applied Covasim to explore the impact of different test-trace-isolate strategies when schools reopened in the UK during autumn 2020 in the absence and presence [20] of requirements to wear masks. Development and implementation details can be found at http://docs.covasim.org with the methodology outlined in [21]. The code used to run the simulations reported in this paper is available from https://github.com/Jasminapg/Covid-19-Analysis/tree/master/7_schools3.

For this study, as in our previous work [19-20], we used Covasim’s default parameters, pre-populated demographic data on population age structures and household sizes and contact patterns in school, community and household setting for the UK, and the population stratified across four population contact network layers: schools, workplaces, households and community settings. The per-contact transmission probability (the risk of transmission during a contact between an infectious individual and a susceptible individual) was assumed to depend on the contact network.

Within the analysis we represent the distinction between primary and secondary schools implicitly via children connected to the schools layer and their interaction in this layer modulated according to their age. An alternative would be to split the schools layer into primary and a secondary schools sub-layers, but this would require a more a more complex way of modelling association between this and adults and other society (e.g. teachers) layers. For the purpose of this analysis, the average effect of our approach is equivalent to the one where there are separate primary and secondary school layers.

### Test, trace and isolation strategies

Covasim accounts for Test, Trace and Isolate strategies as modelled previously [19]. Specifically, testing is incorporated via parameters that determine the probabilities with which people with different symptoms receive a test each day, both for symptomatic and asymptomatic people and based on the reported testing level from https://coronavirus.data.gov.uk. Tracing is quantified by parameters controlling the probability of reaching the contacts of those testing positive, as well as the time taken to reach them. Specifically, we assume that some layers of society would be easier to trace than others (see data sources and calibration section below). Within Covasim we also quantify the level of adherence to isolation as model parameters describing the level of isolation across different layers of the population (see data sources and calibration section below).

### Vaccination strategies

For this study we model a vaccine reducing disease acquisition and transmission with a two-dose regime given 12 weeks apart, to reflect our understanding of vaccination guidelines in place in January 2021 [22]. We model that the full two-dose course reduces the probability of developing symptoms by 95% and per-exposure transmission probability by 30%. Single dose efficacy is assumed to be 70% of the full two-dose course. The second dose is administered 12 weeks after the first dose and with immunity from the vaccination assumed to increase from around 14 days after immunisation with full effect after 21 days after vaccination. Doses are allocated to 200,000 people per day on average, with people 75 or older targeted first and then the roll out following the age prioritisation in the UK.

### Data sources and calibration

Within Covasim’s default contact network generation algorithm, we generated a population of 100,000 agents interacting over the four contact networks layers (households, workplaces, schools, and communities). To reflect the size of the UK population, we used dynamic scaling, as described in detail in [21], of these 100,000 agents up to the population of around 68 million. Dynamic scaling is a standard approach within individual-based-models such as Covasim, that allows for arbitrarily large populations to be modelled whilst maintaining a constant level of precision and manageable computation time throughout and allowing cumulative number of cases to be generated from infection emerging from these agents modelled over time.

Unlike our previous work [19-20] where we modelled one strain (wild type) of SARS-CoV-2 in circulation, here we have modelled the implications of two strains of the virus circulating by simulating a single strain with time-varying infectiousness. Specifically, based on existing findings [4,5] we assume that the new variant is more transmissible. We model this by including a logistic growth function for the relative proportion of increase of the alpha variant from September 1, 2020 to January 31, 2021 such that 30% of infections in December and 90% of infections by the end of January 2021 were caused by the new variant as suggested in [4]. This allowed us to model different levels of infectiousness of B.1.1.7 compared to the wild type, in an approach similar to other work [4].

In the calibration process we fitted the model generated epidemic estimates to the UK epidemic metrics, by performing an automated search for the optimal values of (a) the number of infected people on January 21, 2020, (b) the average per-contact transmission probability across layers, which is then subsequently scaled by layer (c) the daily testing probabilities for symptomatic individuals (*p*_*s*_) over November 2020 -January 2021, and (d) the 3 parameters associated with the modelled logistic curve of the alpha variant strain (number of cases on September 20, 2021, slope of the infectiousness curve and time when saturation is achieved). We used the Optuna optimisation methodology (https://optuna.org) to search the hyperparameters space for optimal values of these parameters that minimised the sum of squared differences between the model’s estimates of cumulative diagnoses, cumulative deaths and cumulative hospital admissions, and data on these same three indicators between January 21, 2020 and January 25, 2021 collated from the UK government’s COVID-19 dashboard (https://coronavirus.data.gov.uk). These six particular model parameters were selected as the most important to estimate because of the considerable uncertainties around them. The calibrated model was able to reproduce the UK epidemic trajectory between January 21, 2020 and January 25, 2021 (Supplementary Figure S1).

Within the model we also simulated the effect of the two national lockdowns that were imposed in 2020. For the first national lockdown, when schools closed and as in [19,20], we modelled a 98% reduction in the per-contact transmission probabilities from March 23, 2020 within schools and an 80% reduction in transmission within workplace and community settings, and increased these in a phased way since the phased relaxing of the lockdown measures from June 1, 2020. For the second lockdown, between November 5, 2020 and December 3, 2020 during which schools remained open, we assumed a reduction in the per-contact transmission probabilities by 37% in schools i.e. simulated 63% of transmission within schools remaining from September. This was modelled as aggregated reduction in transmission due to hygiene, mask usage and other social distancing measures in place within schools to reduce transmission, and as described in details in [20]. Furthermore, since January 2021, children of key workers have been attending school, with estimates suggesting that around 1 in 5 primary school students and 5% of secondary schools students attending – on average 14% of children in attendance [18]. We included this in the modelling framework simulating 14% transmission in schools since January 4, 2021.

The level of reduction in transmission in households, workplaces and community that we modelled was informed by Google mobility data [23]. Specifically, for the household transmission we modelled increased transmission since November 2020 of 25% and in line with the average monthly level of increased household mobility in the Google data [23]. For the workplaces and community, during the November lockdown, we also used Google mobility data to obtain a broad range of the change, but we also needed to adjust these during the calibration process. Specifically, we simulated workplace and community transmission to be reduced by 80% and 80% of their pre-COVID-19 levels during the first lockdowns, and 70% and 60% during the second lockdown.

We also used publicly available weekly data from NHS Test and Trace to estimate the rate of tracing of contacts of those testing positive since the start of the programme on May 28, 2020 [24]. For each weekly period, we collated the percentage of people testing positive who were interviewed, the percentage of those reporting contacts and the percentage of contacts traced. We used these percentages to produce an overall estimate of the percentage of contacts of those tested positive who were traced. We then computed the monthly average effective contact tracing level and in our previous work, we used this data to produce a monthly effective contact tracing level. As an extension, here we additionally assumed that tracing levels differ depending on the type of contact, and assumed that 100% of household contacts can be traced within the same day, 50% of school and workplaces can be traced within one day and 10% of community contacts can successfully be traced within 2 days; giving an average of 53% of contacts traced across different layers and comparable with reported monthly values from [24]. We also assumed that asymptomatic testing is available across all society layers and modelled this in line with reported numbers in the UK (0.076% May-August 2020, 0.28% August-October 2020 and 0.63% since November 2020 from https://ourworldindata.org/coronavirus-testing).

### Scenarios

We modelled five different scenarios as postulated reductions in transmission within schools, community and workplaces, which are shown in Table 1 and briefly summarised below. For each scenario we predicted the number of new daily cases, cumulative deaths and R until April 30, 2021.

**Table 1:**
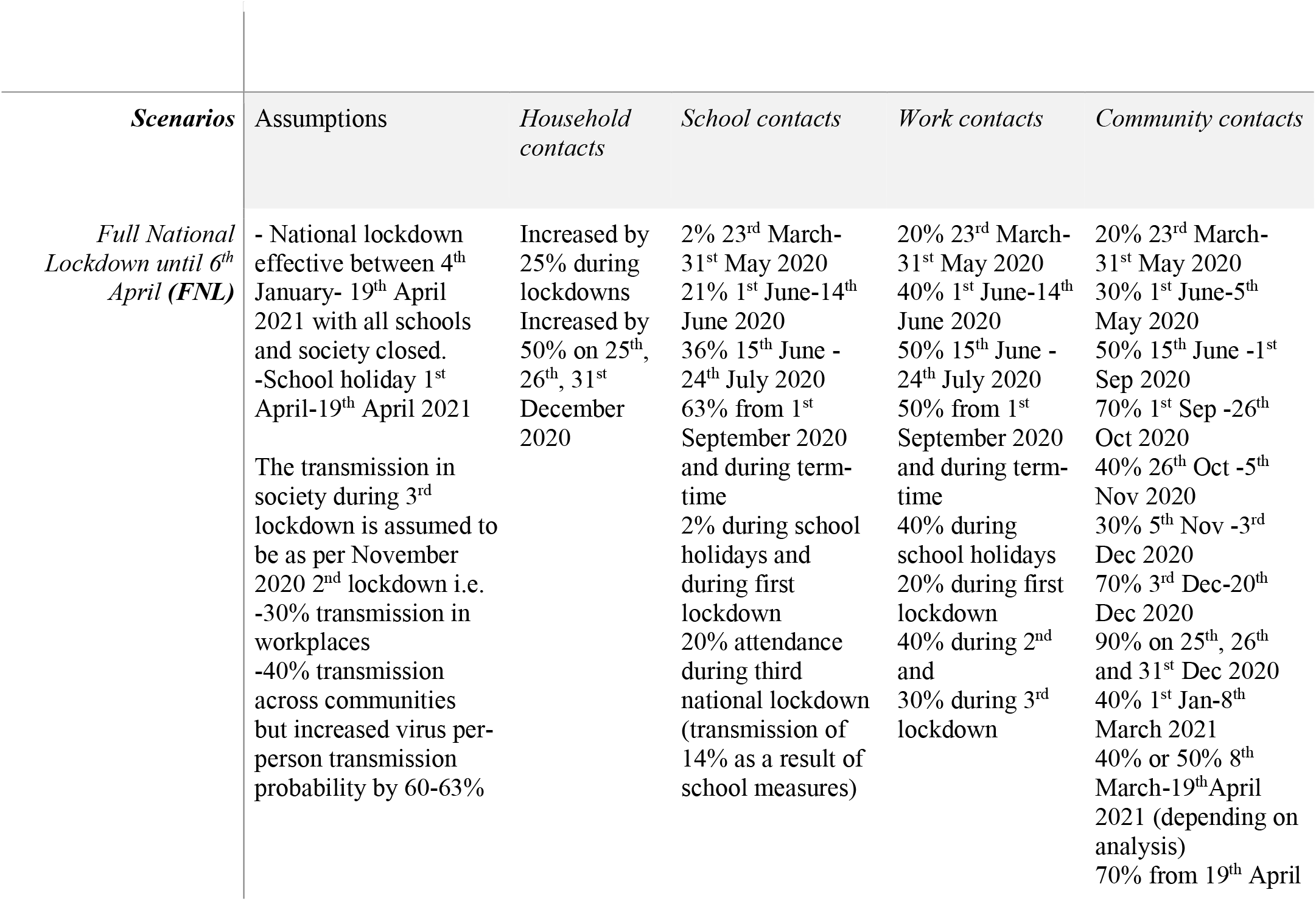
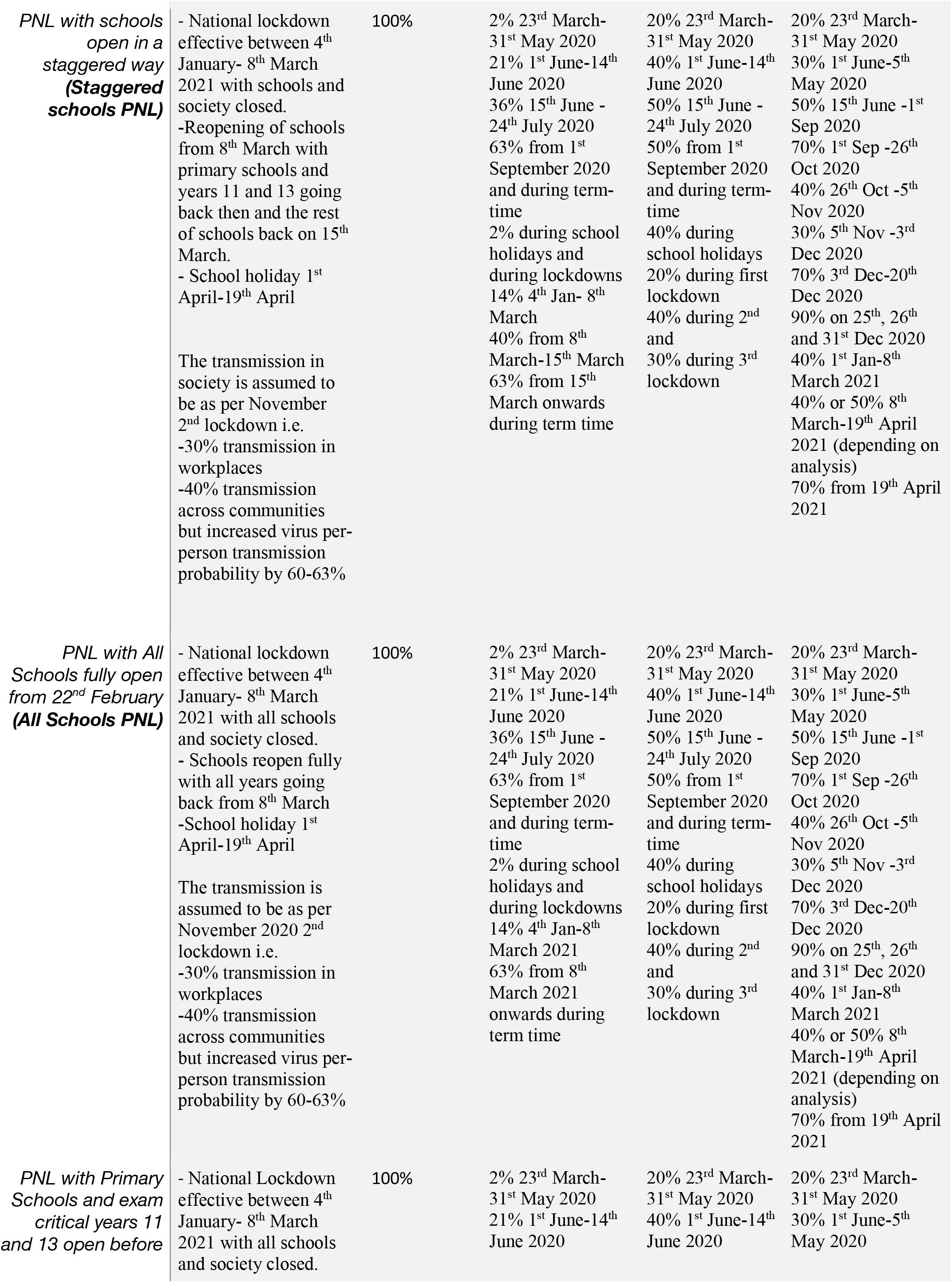
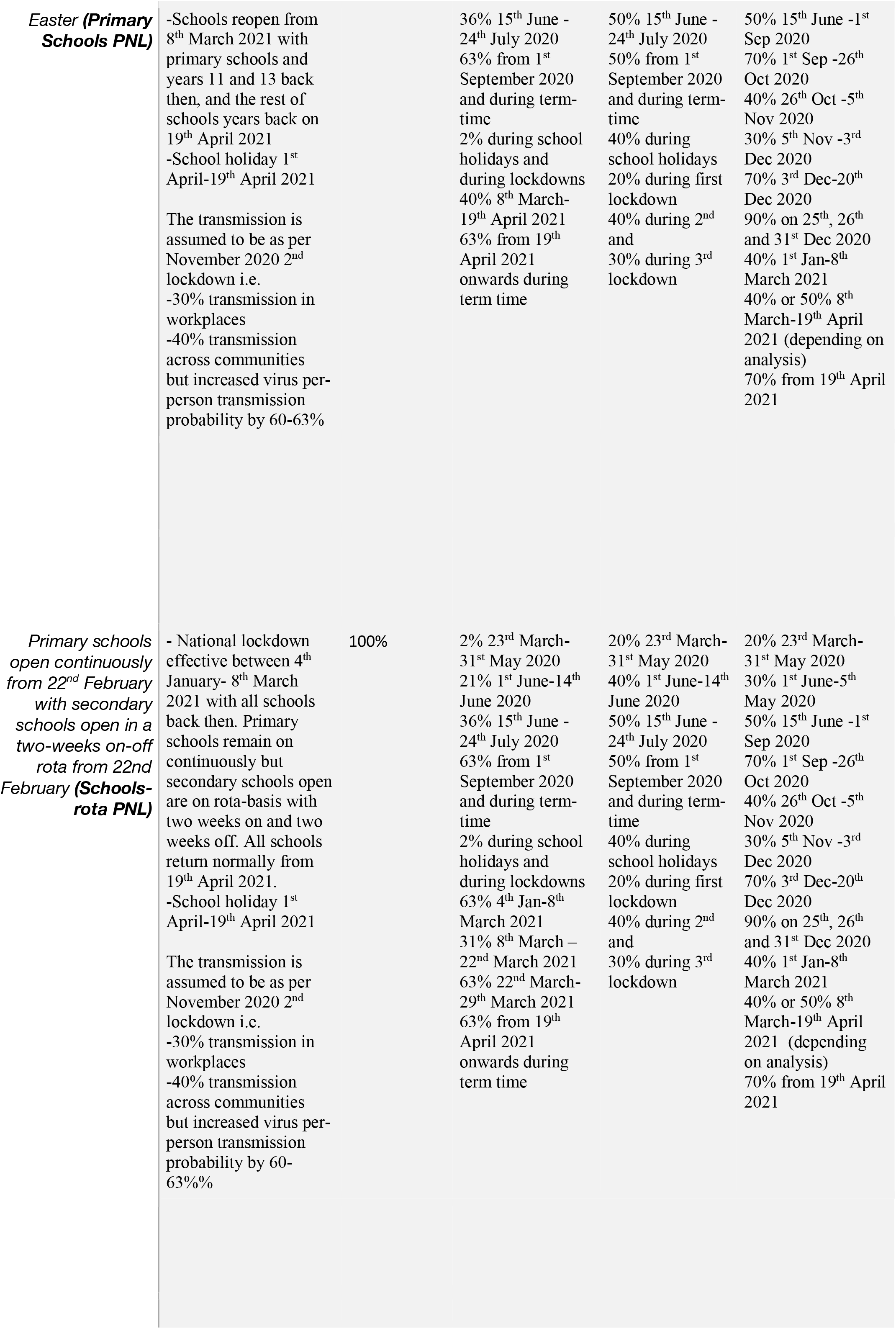
Scale factors applied to daily SARS-CoV-2 transmission probabilities in households, schools, workplaces, and the community under the three simulated scenarios

Scenario 1: **FNL** between January 4 and April 19, 2021, with all schools and reduction in transmission within workplaces, homes and community modelled as in the November 2020 lockdown until April 19.

Scenario 2: **Staggered PNL**: FNL between January 4 and March 8, 2021 with PNL after March 8, 2021 with all schools opening in a staggered way: primary schools and years 11 and 13 of secondary schools from March 8 and the rest of secondary school years from March 15. A reduction in transmission within workplaces, homes and community is modelled as in the November 2020 lockdown until April 19.

Scenario 3: **Full-return PNL**: FNL between January 4 and March 8, 2021 with PNL after March 8, 2021 with all schools opening from March 8. A reduction in transmission within workplaces, homes and community is modelled as in the November 2020 lockdown until April 19.

Scenario 4: **Primary-only PNL**: FNL between January 4 and March 8, 2021 with PNL after March 8, with only primary schools and exam critical years (years 11 and 13) opening on March 8 and the rest of the secondary schools opening on April 19 when society also reopens. A reduction in transmission within workplaces, homes and community is modelled as in the November 2020 lockdown until April 19.

Scenario 5: **Part-Rota PNL**: FNL between January 4 and March 8, 2021 with PNL after March 8 with all schools opening from March 8 with primary schools remaining continuously open but secondary schools open on a two-weeks rota system until April 19 when schools and society reopen. A reduction in transmission within workplaces, homes and community is modelled as in the November 2020 lockdown until April 19.

### Sensitivity analyses

Although evidence is emerging on the relative susceptibility to the virus for children compared to adults [25], there is a uncertainty around the exact numbers. To account for this, in the main analysis we assume that primary school children (0-10 years old) are 50% less susceptible than secondary school children (11-18 years old) or adults (>18 years old), with these two latter groups having the same susceptibility [25]. The sensitivity analysis then explored whether the results changed if all age groups have the same susceptibility as adults.

There is also some contention [26] regarding the degree to which transmission in society increases as a result of schools reopening. Our main scenarios assume that it would not, but we also conduct additional sensitivity analyses to explore how the results would change if community transmission increased as a result of reopening of schools.

## Results

Our calibration across different scenarios of children’s susceptibility between 50-100% of that of adults’, is consistent with the new alpha variant being around 60% more transmissible than the wild type of SARS-CoV-2.

The separate panels in Figure 2 report the epidemic projections of daily new COVID-19 cases, cumulative deaths and R (as rows) across the two scenarios (as columns) until April 30, 2021. Projections for the daily infections and total infections for each scenario over the period from February 22 and April 30, 2021 are shown in Figure 3(a)-(b).

**Figure 2:**
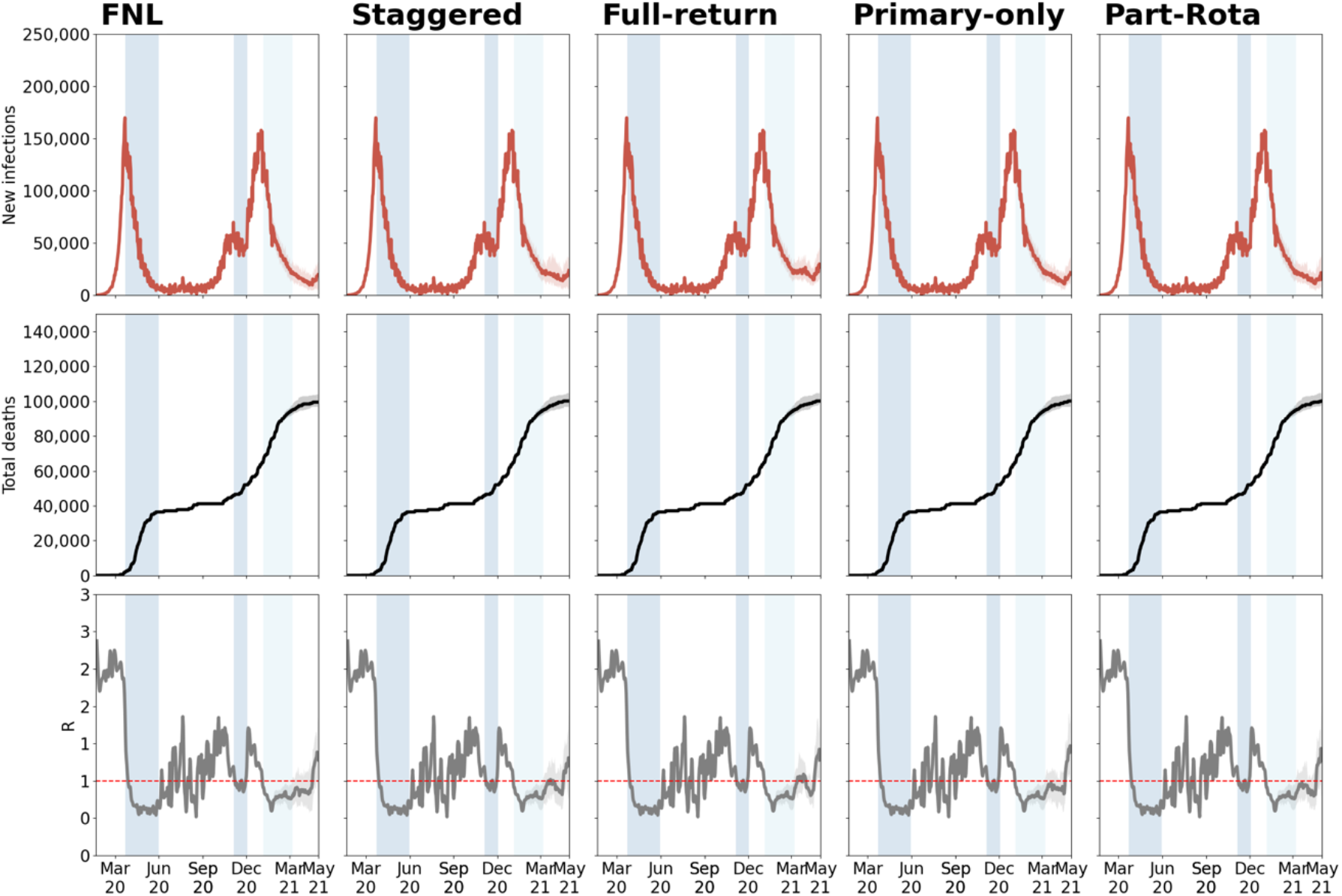
Model-predicted epidemic trajectories of the calibrated model until April 30, 2021 across FNL and PNL with different reopening strategies and under the assumption that susceptibility in 0-10 years old is 50% less than across other ages and that community transmission remains the same as in November lockdown between March 8,2021 and April 19, 2021. Medians across 30 simulations are indicated by solid lines and the 25% and 75% quantiles by shading. The blue bands represent the past and current national lockdowns.

**Figure 3:**
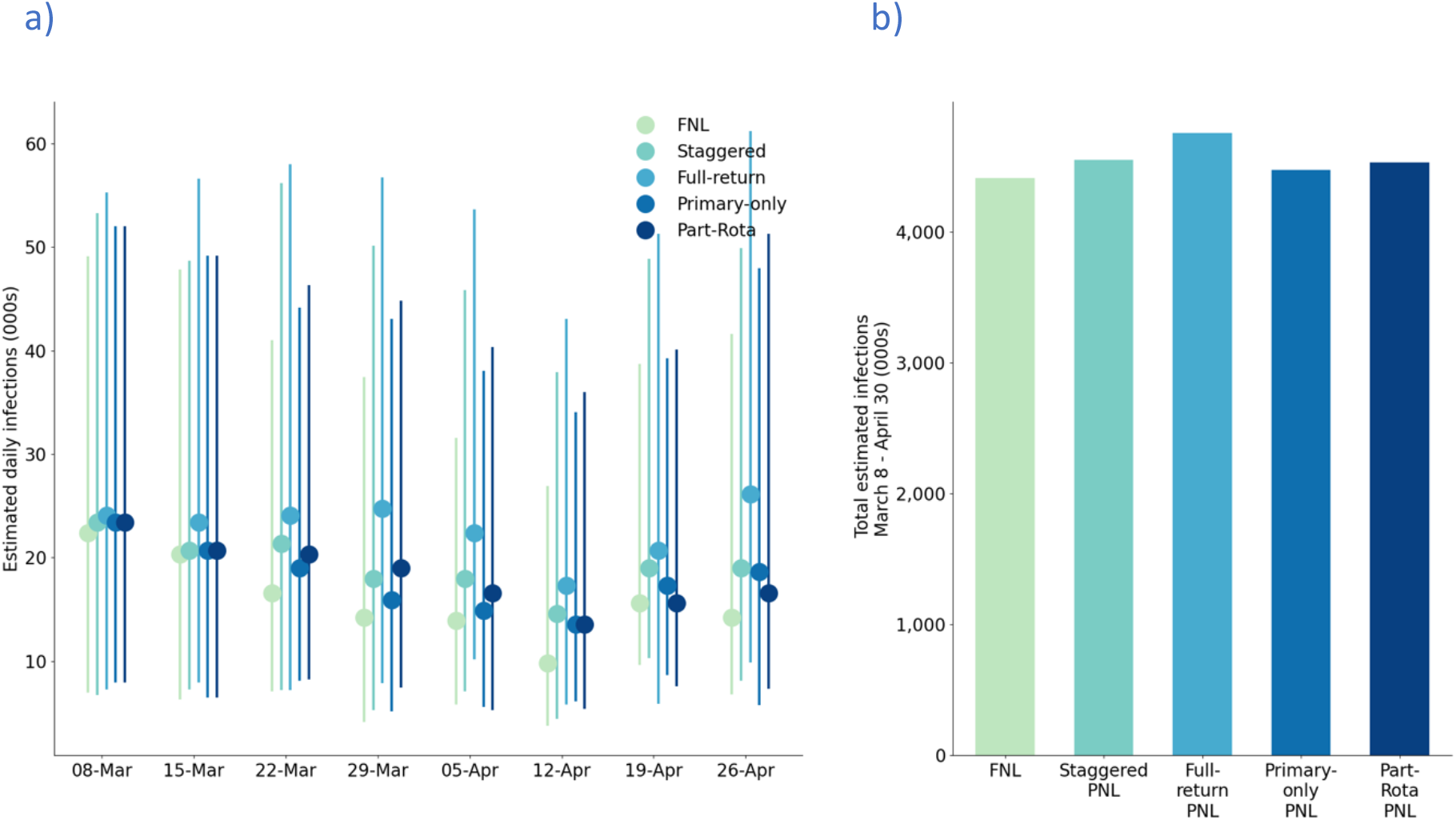
Model predicted estimated daily (a) and total (b) infections in the period from February 22 to April 20, 2021 from the calibrated model across the five different scenarios: FNL, Staggered PNL, Full-return PNL, Primary-only PNL and Part-Rota PNL under the assumption that susceptibility in 0-10 years old is 50% less than across other ages and that community transmission remains the same as in November lockdown between March 8,2021 and April 19, 2021. In (a) we show the point estimates as well as the uncertainty range around this for each scenario.

Our results suggest that strict social distancing measures, i.e. national lockdown, were required to contain the spread of the virus and control the hospitalisations and deaths during January and February 2021. We estimate that the national lockdown reduced the number of cases by early March to a similar level as in October with R also falling and remaining below 1.

We find that, across each scenario, the number of new infections would have been expected to decrease over January and early February lockdowns. This decrease is more sustained in the FNL than any of the PNL scenarios. Impacts upon deaths are lagged, with plateauing of cumulative deaths seen from February in each scenario. This is due to the vaccine effect having been modelled with a delay of 21 days. Overall, we predict a rise in the number of infections and increase in R following the reopening of schools, and a possible shift in R above 1 once society opens also.

Reopening primary schools and exam critical years with the rest of secondary schools to reopen later, would result in the least increase in both infections and R over the study period. The opening of secondary schools would result in a larger rise in the number of infections and the R value rising to close to 1 in early March. The rise in cases was estimated to be most rapid for the Staggered PNL and Full-return PNL. The Part-Rota PNL, where primary schools remain open while secondary schools are open for two-weeks and closed, with online teaching provided, for two-weeks was also associated with an increase in cases once secondary schools open, but not sufficient to push R above 1. The increase in cases that pushes R above 1 occurs in each scenario once the rest of the society opens up on April 19, 2021.

### Impact of FNL

The FNL scenario reduced R below 1 and markedly reduced new infection rates from mid-January, thus acting as an effective short-term measure for suppression of COVID-19 infections. But once the lockdown was relaxed, with some form of school reopening from March 8, our projections indicated that some increase in cases and R would follow, especially following the reopening of the rest of society, simulated here from April 19, 2021.

### Impact of PNL with staggered or full schools reopening

If, instead of continuing the 3^rd^ national lockdown under our FNL scenario, schools were to reopen from March 8, 2021 either fully or in a staggered way with primary schools and years 11 and 13 returning from March 8, 2021 and the rest of secondary school students from March 15, 2021, we estimated that there would be an increase in new infections during March and April compared with a FNL and an increase of R to closer to 1. These scenarios would result in a higher predicted level of new infections than a FNL from the end of lockdown in February. Furthermore, fully-reopened schools or staggered PNL would result in the highest numbers of daily and cumulative cases in March and April compared to other scenarios, although confidence intervals overlap (Figure 3(a)-(b)).

### Impact of PNL with primary schools open

Reopening of primary schools from March 8, 2021 with either two-weekly rota system for secondary schools or a delayed return to school for secondary school students, with secondary schools returning to face-to-face learning until April 19, 2021 would result in the least increase in new infections during March and April compared to other scenarios (Figure 3(a)) and a lesser increase in R (Figure 4).

**Figure 4:**
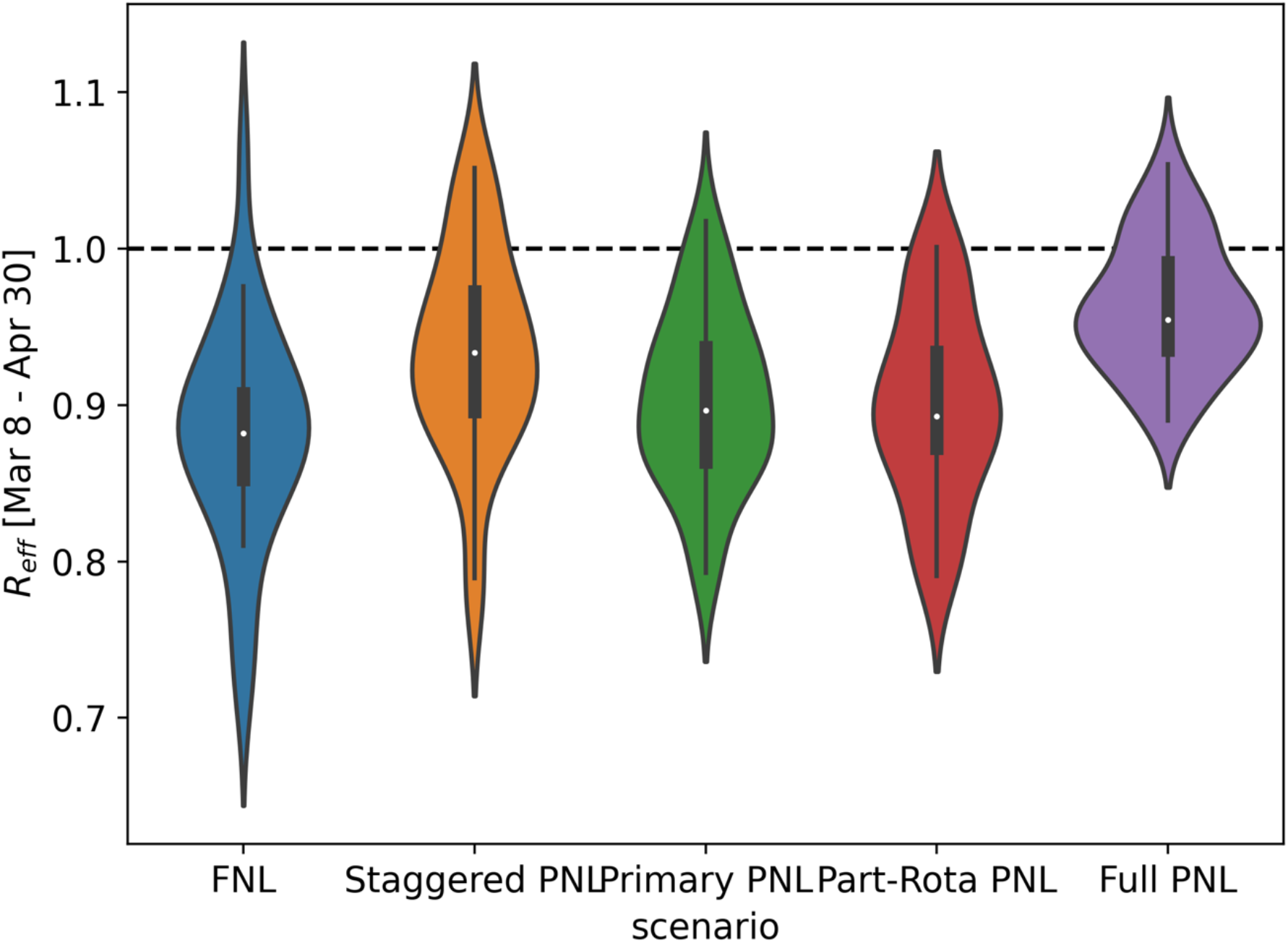
Estimated values of the mean effective reproduction number between March 8, 2021 and April 19, 2021 under the current epidemic trajectory and with different assumptions of reopening schools from March 8, 2011. We show the point estimate and the uncertainty range for each scenario.

## Discussion

Our calibration is consistent with the new alpha variant being around 60% more infectious in line with existing literature [4,5]. The model estimates there were approximately 1.2 million active infections across the UK as of January 16, 2021 implying that 1 in 60 people in the UK were infected with COVID-19 at that time. This is highly similar to ONS estimates for January 16, 2021 suggesting there were 1,149,300 (95% credible interval: 1,082,300 to 1,220,110) active infections and approximately 1 in 59 (95% credible interval: 1 in 62 to 1 in 55) [27] infected across the UK. We also estimated that R may be around 0.8 by March 8, 2021.

Our modelling suggests that cases and R would have been lowest had the FNL continued until April 19, 2021, compared to any of the scenarios with some schools reopened. Reopening primary schools and exam critical years only, or having primary schools open continuously with secondary schools on a two-weeks on-off rota, would have led to a lower increase in cases and R than a scenario under which all schools were opened.

In all scenarios, the reopening of schools together with relaxation of broader lockdown of society would lead to a rise in infection rates and rise in R above 1 from April 19, 2021 onwards. Hence it is important to effectively roll out a mass vaccination strategy during lockdowns; we are undertaking further work to examine different vaccine dosing strategies and the trade-off between the speed and the efficacy of different vaccination regimes. This is important for decisions around future mitigation of the virus resurgence once lockdowns are lifted.

Given the harms of closing schools, it is important to open them as soon as it is safe to do so, and to enact sufficient countermeasures to minimise the risks of resurgence of infections and deaths and overwhelming hospitals. Our findings, conducted in February of 2021, suggested that reopening of schools under different scenarios from March 8, 2021 could plausibly have been consistent with maintaining R below 1 even without factoring in the impact of increases in vaccination rates or increased testing and countermeasures within schools. Thus, we find that reopening of schools at this time was a potentially valid policy option, and one which could be further strengthened by the incorporation of additional strategies within schools and society, including further expansion of on-site testing at schools, testing at the earliest onset of symptoms, wider use of face coverings within schools, vaccinating teachers as a priority group as well as achieving high vaccine coverage and maintain social distancing in other sectors. Many of these strategies have been modelled using Covasim in other work [33]; for example, within the Seattle context it was found that daily symptom screening would reduce the average outbreak size within schools to below 5 even with high prevalence in the community, compared to ∼15 without any daily symptom screening.

We note that reopening schools when prevalence remains high in the general population may lead to an increased risk of COVID-19 transmission within schools and therefore higher cases in children and adolescents. However severe COVID-19 disease and post-inflammatory complications remain extremely rare in these age-groups [31-32].

As with any modelling study, we have made a number of assumptions when simulating the epidemic. Firstly, as in our previous work [19-20], we assumed that symptomatic infections account for 70% of onward-transmitted infections, in line with other published work [28-30]. Secondly, we have modelled the presence of the alpha variant by simulating time-varying infectiousness for the virus strains from September 2020. Different virus strains, emerging from mutations, can have different characteristics; one of the key ones is their transmissibility (or infectiousness) and severity. In this study, based on recent reports [4,5] we modelled infectiousness as the difference between the two viral strains. Simply speaking we fitted different infectiousness values for each strain, assuming that the growth rate for these strains and also that one virus strain dominates the spread within each epidemic wave. Under this assumption, we modelled the contribution from the dominating strain to induce larger infectiousness. We also note that in this work we did not assume that the alpha variant had higher severity than the wild type as there is uncertainty around this. We are undertaking follow on work that will explore the transmissibility and severity parameter space across variants in more detail. Thirdly, we did not use regional variations in cases, R value or the proportion of alpha presence. With these assumptions on uniformity of the virus spread, and the hypothesis on the two strains, we note that we may underestimate the number of new infections because we don’t explicitly model how much the new variant predominate and how across different regions. It is possible that the future spread across different regions may lead to increased net transmissibility. In a follow up study, we will focus on exploring this for London specifically - one of the hotspots at the onset of the second epidemic wave. Additionally, our follow on work on the impact of reopening after the second wave will also consider the impact of reopening of schools and other parts of society at different stages/times on hospital admissions, intensive care units capacity and deaths. The balance between reopening society with increased social mixing, the impact from continual vaccination roll-out on hospitalisations and deaths as well as the reduction of onward transmission, and the possibility of emerging of new more transmissible or vaccine-escapee variants will be increasing important later in 2021 and beyond. Our ongoing and future work will look at exploring these whilst tracking the epidemic status in the UK.

In future we will also look at more granular distribution of cases, hospitalisations and deaths across different ages and explore whether reopening schools under different scenarios leads to rise in infections in school age children proportional to their representation in society [34]. Furthermore, looking at age mixing/chains of transmission is an important extension of our work to consider in future, as a single infection in a school-age child will not stay in that age group only but propagate through society levels.

Unlike in our previous work [19-20] where the level of contact tracing to be the same across all contact network layers, in this work we used different tracing levels across layers to better represent the contact tracing methodology. We have assumed that tracing within households will be 100% effective within the same day, while within schools and workplaces it will be 50% effective within one day. i.e. will miss about half of the cases within one day. We acknowledge that this may be an underestimation if effective cohorting is in place, with NHS Test and Trace reporting that different fraction of contacts can be reached in complex (e.g. schools and workplaces) and non-complex cases (e.g. households). This suggests that cases have better tracing in institutions where contacts identities are better known. The fraction of contacts reached within the community are not reported and we expect that challenges associated with finding, tracing and isolating are with contacts in the community; within the model we have assumed that only 10% of the community contacts are traced within two days. With these assumptions across layers, the average tracing level is around 53% which is agreeable with recent reported values [24].

As with any stochastic modelling, there is uncertainty in our predictions, most notably in the confidence intervals in Figure 3(a). Our results are based on taking median simulation from a stochastic process with a degree of uncertainty may increase when making longer time future predictions. Hence, in Figures 2-3 we only project for 8 weeks into the future from March 8, 2021 and note that projecting results of any model, including ours, too far into the future based on current data is unwise due to the high level of uncertainty. Importantly, estimation of future epidemic trajectories will depend on the effect of the continual roll-out of mass vaccination against COVID-19 and further additions to it.

Finally, we note that these results were produced for the particular epidemiological context of the UK over the period January 2021-April 2021, and will not necessarily be easily generalisable except to other settings that have similar contact patterns both within and outside schools, mobility restriction policies, and seroprevalence levels. These choices were made since our aim was to produce not just a theoretical study, but one with real-world implications for the UK COVID-19 epidemic over the study period.

In summary, our analyses ran in February 2021, offer evidence that the national lockdown imposed from January 2021 with schools closed was likely to have been successful in suppressing the wave of COVID-19 cases that emerged towards the end of 2020. We also provide evidence that continued epidemic control was achievable even with cautious reopening of schools from March 8, 2021 whilst continuing the vaccination efforts initiated from December 2020.

## Data Availability

All data and numerical code used in this analysis are available at https://github.com/Jasminapg/Covid-19-Analysis

## Contribution

JPG and RV came up with the idea of the study. JPG, RMS and KR developed the specific modelling framework, based on the Covasim model developed by CCK, RMS, DM and DJK. CCK, RMS, DM, DJK, KR and JPG collated data for the parameters used. JPG ran the modelling analysis with input from RMS and KR. JPG, RV and CB defined the different scenarios in the UK context following conversations with Scientific Pandemic Influenza Modelling Group (SPI-M) and Scientific Pandemic Influenza Behaviour Group (SPI-B) which give expert advice to the UK Department of Health and Social Care and wider UK Government. JPG wrote the manuscript with input from RMS, CB, RV, CCK, KR, WW, DM and DJK. All authors approved the final version. JPG is the manuscript’s guarantor.

## Statement on data quality

All data and numerical code used in this analysis are available at https://github.com/Jasminapg/Covid-19-Analysis/tree/master/7_schools3

