## Supplementary material for "Modelling the impact of reopening schools in the UK in early 2021 in the presence of the alpha variant and with roll-out of vaccination against SARS-CoV-2" (from another version of this preprint: version 1, 2021-02-09)

#### Incorporating the SARS-CoV-2 new variant B.1.1.7 in the model

Following existing reports that the new variant is more transmissible, we model the new strain with an increase in proportion from September 1, 2020 to January 31, 2021 following a logistic growth function with the % of infections by end of January caused by the new variant fitted during the calibration. This allowed us to model different levels of infectiousness of the B.1.1.7 compared to SARS-CoV-2, in an approach similar to recent work in [5].

#### Model calibration

| **Calibration of the model** | **Susceptibility of 0-10 years old is 50% of that in other age groups** | **Susceptibility is the same across all ages** |
|---|---|---|
| Initial seeding of infectious persons on 21/01/2020 | 1500 | 1500 |
| Infectiousness of SARS-CoV-2 ($\beta$) | 0.00815 | 0.0078 |
| Infectiousness of B.1.1.7 | 60% | 63% |
| % of infectious due to B.1.1.7 at end of January | 62% | 75% |
| $p_s$ in March | 0.009 | 0.009 |
| $p_s$ in April | 0.009 | 0.009 |
| $p_s$ in May | 0.012 | 0.012 |
| $p_s$ in June and July | 0.0277 | 0.0277 |
| $p_s$ August | 0.0377 | 0.0377 |
| $p_s$ in September, October, November and December | 0.0877 | 0.0877 |
| $p_s$ in July and August | 0.0277 | 0.0277 |
| $p_{as}$ in May, June, July and August | 0.00075 | 0.00075 |
| $p_{as}$ in September, October, November and December | 0.02 | 0.02 |
| Tracing level in June | 42% | 42% |
| Tracing level in July | 47% | 47% |
| Tracing level in August | 44% | 44% |
| Tracing level in September-December | 50% | 50% |
| Adherence to isolation in May-July (%) | 90% | 90% |
| Adherence to isolation in August (%) | 30% | 30% |
| Adherence to isolation in September-December (%) | 50% | 50% |

**Table S1**: Parameters fitted during the calibration across both scenarios. $p_s$ is the daily testing probabilities for symptomatic individuals while ($p_{as}$) is the daily testing probabilities for asymptomatic individuals.

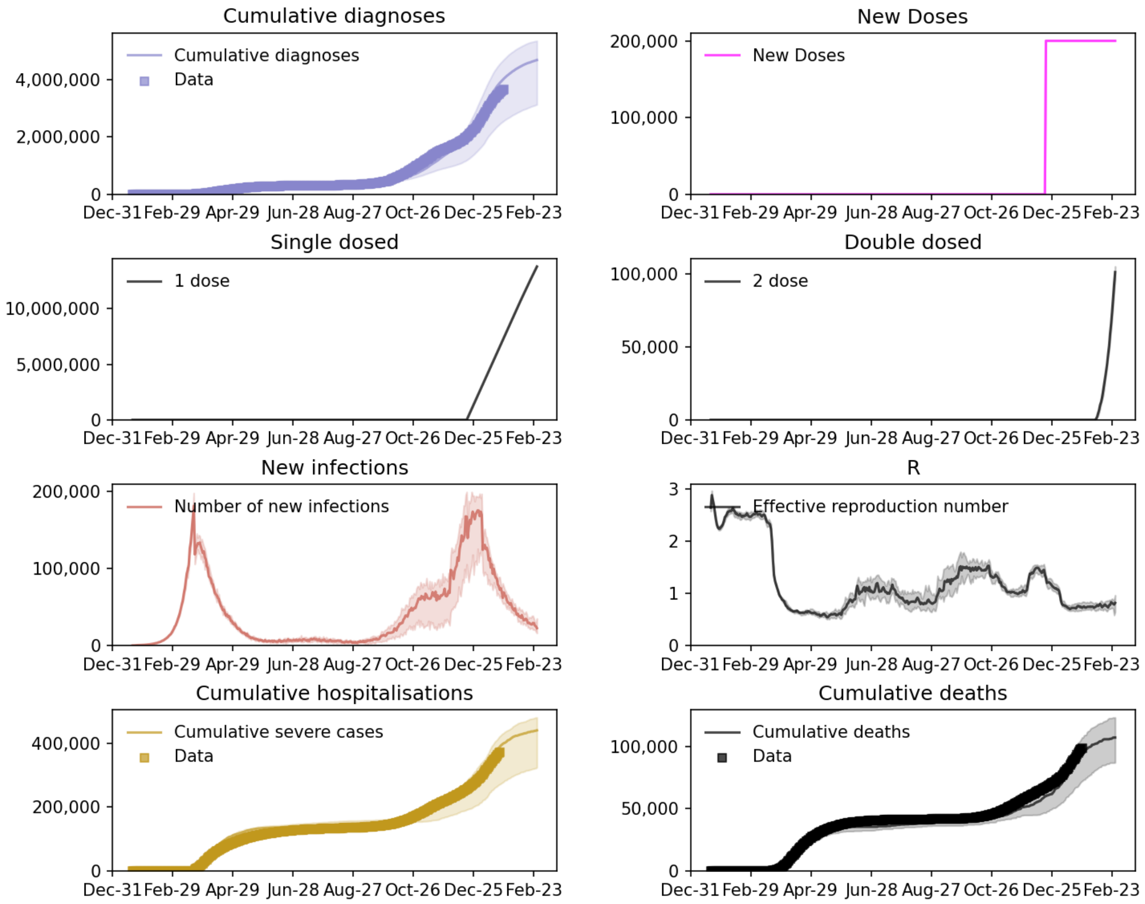


**Figure S1:** Results of the model calibration for the main scenario showing the matching of the model projected cumulative COVID-19 cases, cumulative deaths and admissions to hospital associated with COVID-19 with the data from https://coronavirus.data.gov.uk between 21/02/2020-25/01/2021. We also show the number of new infections, effective reproduction number R and the cumulative first and second dose of the vaccination in the model. Data are shown in thick black/blue/yellow lines, medians across thirty simulations are indicated by thin lines and 25% and 75% quantiles by shading.

### Sensitivity analysis 1: Impact of increased community transmission with schools open

This sensitivity analysis explored whether the main results would change under the scenario that community transmission increases by 10% with reopening of schools. This is based on assuming that reopening schools implies larger mixing in community traveling to and from schools via public transport with parents (primary-school children) or individually (secondary school children). Figure S4-S7 are equivalent to Figures 1-3 from the main manuscript.

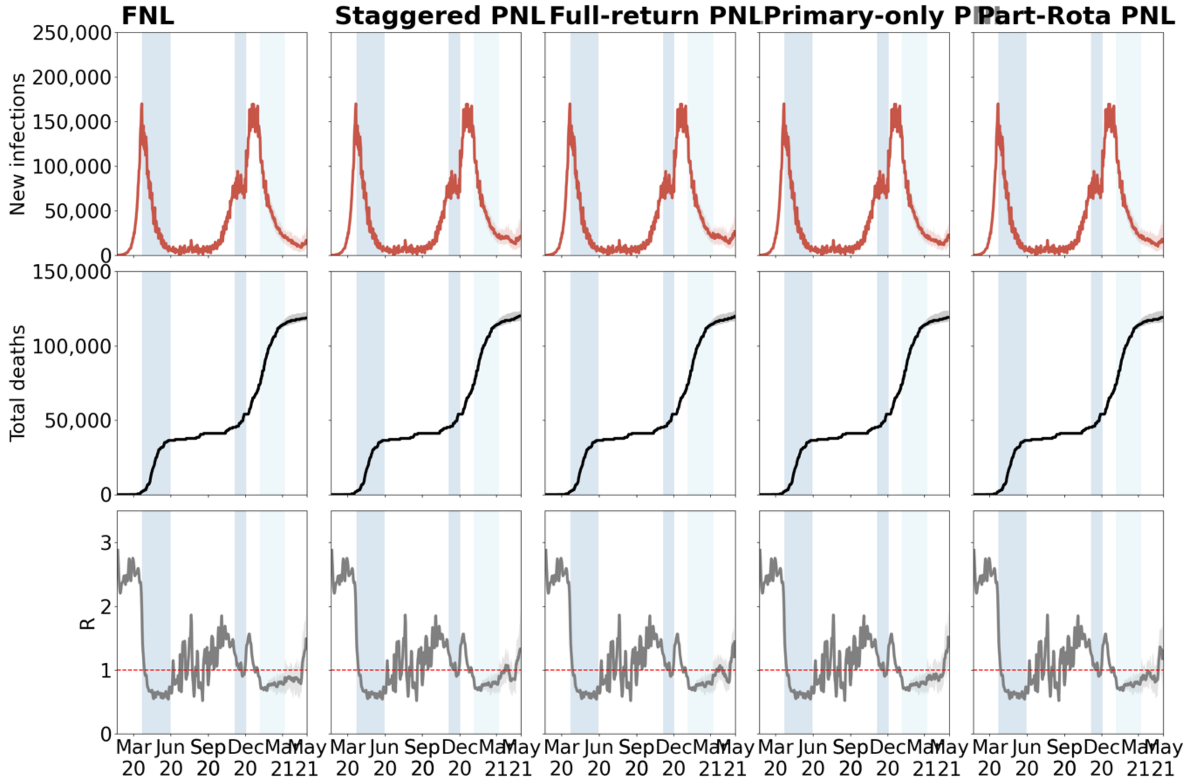


**Figure S2:** Model-predicted epidemic trajectories of the calibrated model until April 30, 2021 across FNL and PNL with different reopening strategies and under the assumption that all ages have same susceptibility but community transmission is 10% higher than during November lockdown between March 8,2021 and April 19, 2021. Medians across 30 simulations are indicated by solid lines and the 10% and 90% quantiles by shading. The blue bands represent the past and current national lockdowns.

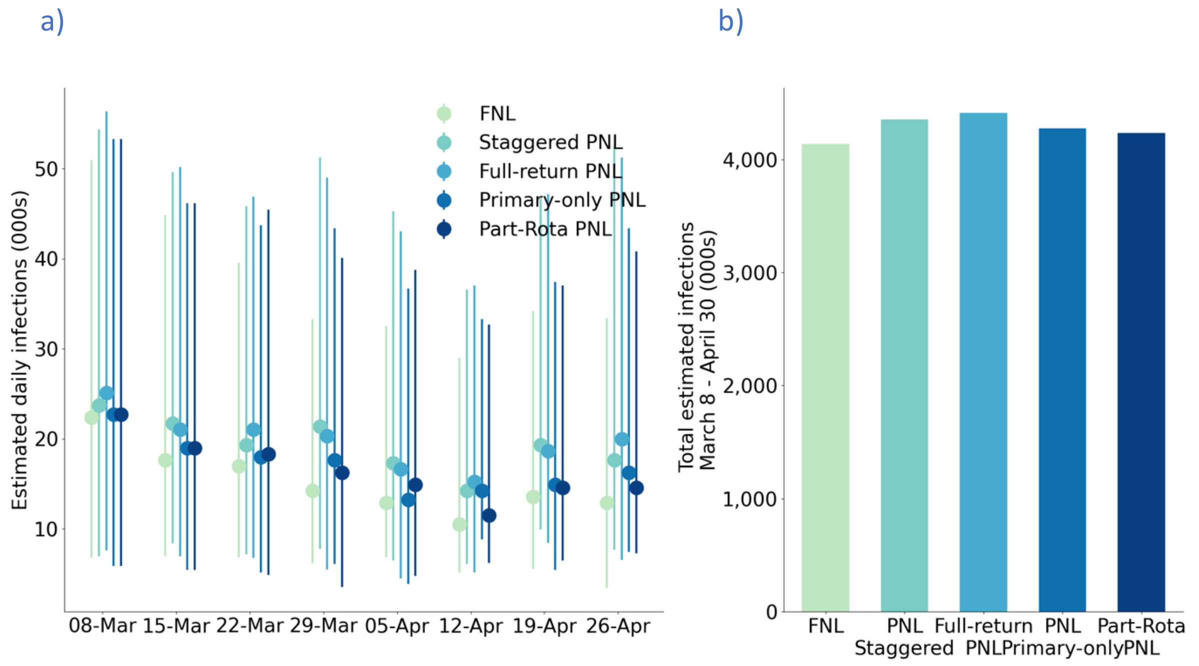


**Figure S3:** Model predicted estimated daily (a) and total (b) infections in the period from March 8 to April 30, 2021 from the calibrated model across the five different scenarios: FNL,

Staggered PNL, Full-return PNL, Primary-only PNL and Part-Rota PNL under the assumption that susceptibility in 0-10 years old is 50% less than across other ages and that community transmission is 10% higher between March 8,2021 and April 19, 2021 than during the November lockdown. In (a) we show the point estimates as well as the uncertainty range around this for each scenario.

### Sensitivity analysis 2: Susceptibility of all age groups is the same

For this sensitivity analysis we repeated the simulations from the main analysis but under the assumption that susceptibility of all ages is the same and explored future epidemic projections if community transmission remains the same as in November lockdown between March 8,2021 and April 19, 2021 or increases by 10% when schools reopen in some way. The calibration required per-person transmission probability $\beta$ to be reduced to 0.0078 and we needed to adjust parameters that described the logistic growth of B.1.1.7 as per Table S1. The calibration outcome is shown in Figure S2.

Figures S3-S4 are equivalent to Figures 1-3 from the main manuscript and contain the results in the scenario where that susceptibility of all ages is the same and community transmission remains the same as in November lockdown between March 8,2021 and April 19, 2021.

Figures S5-S6 are equivalent to Figures 1-3 from the main manuscript and contain the results in the scenario where that susceptibility of all ages is the same but community transmission between March 8,2021 and April 19, 2021 is 10% higher than that of the November lockdown.

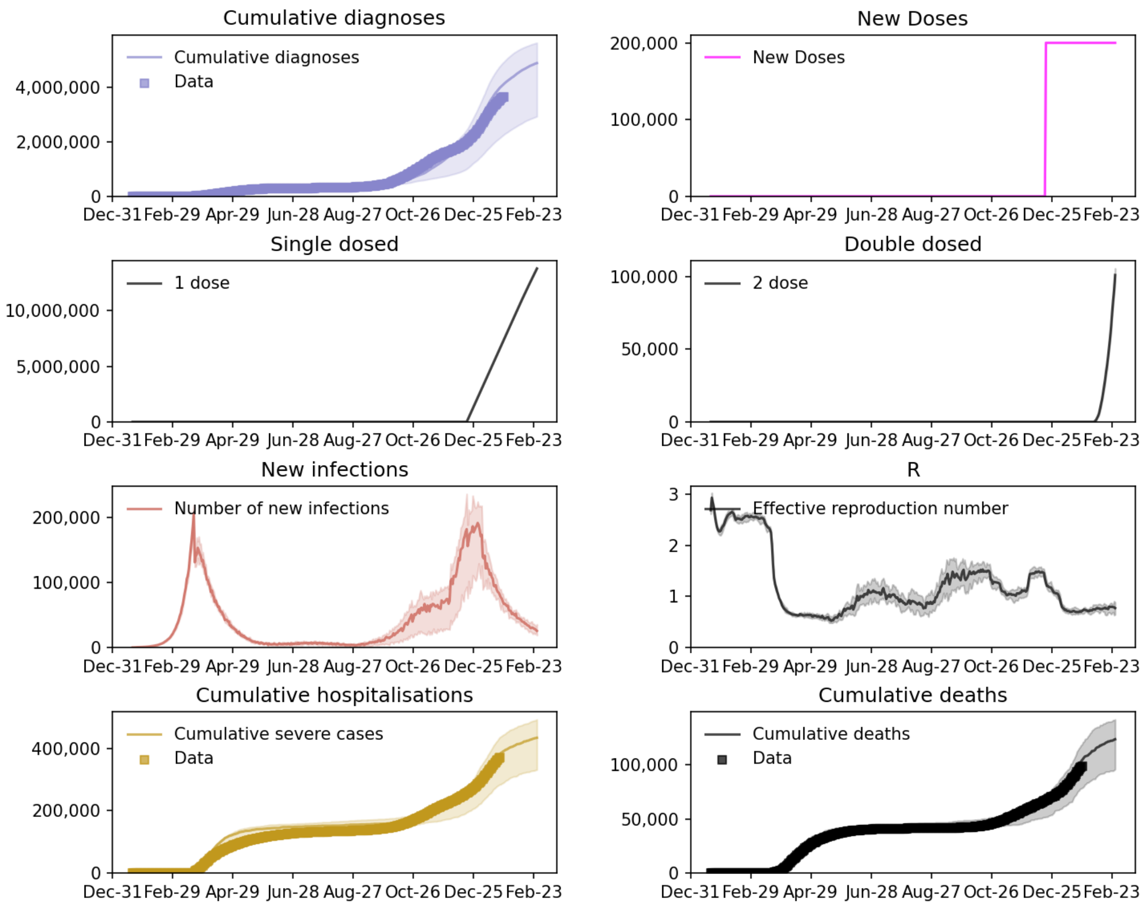


**Figure S4:** Results of the model calibration for the main scenario showing the matching of the model projected cumulative COVID-19 cases, cumulative deaths and admissions to hospital associated with COVID-19 with the data from https://coronavirus.data.gov.uk between 21/02/2020-25/01/2021. We also show the number of new infections, effective reproduction number R and the cumulative first and second dose of the vaccination in the model. Data are shown in thick black/blue/yellow lines, medians across thirty simulations are indicated by thin lines and 25% and 75% quantiles by shading.

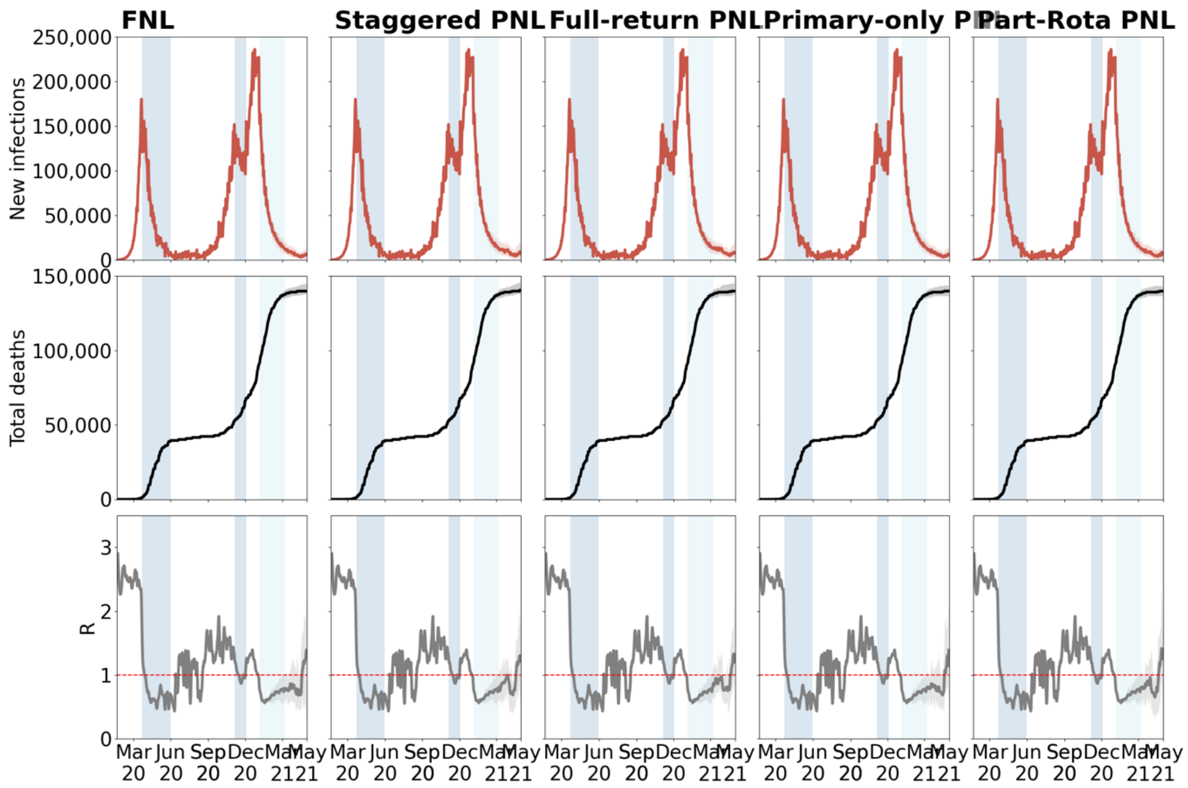


**Figure S5:** Model-predicted epidemic trajectories of the calibrated model until April 30, 2021 across FNL and PNL with different reopening strategies and under the assumption that susceptibility of all ages is the same and that community transmission remains the same as in November lockdown between March 8,2021 and April 19, 2021. Medians across thirty simulations are indicated by solid lines and the 25% and 75% quantiles by shading. The blue bands represent the past and current national lockdowns

a)

b)

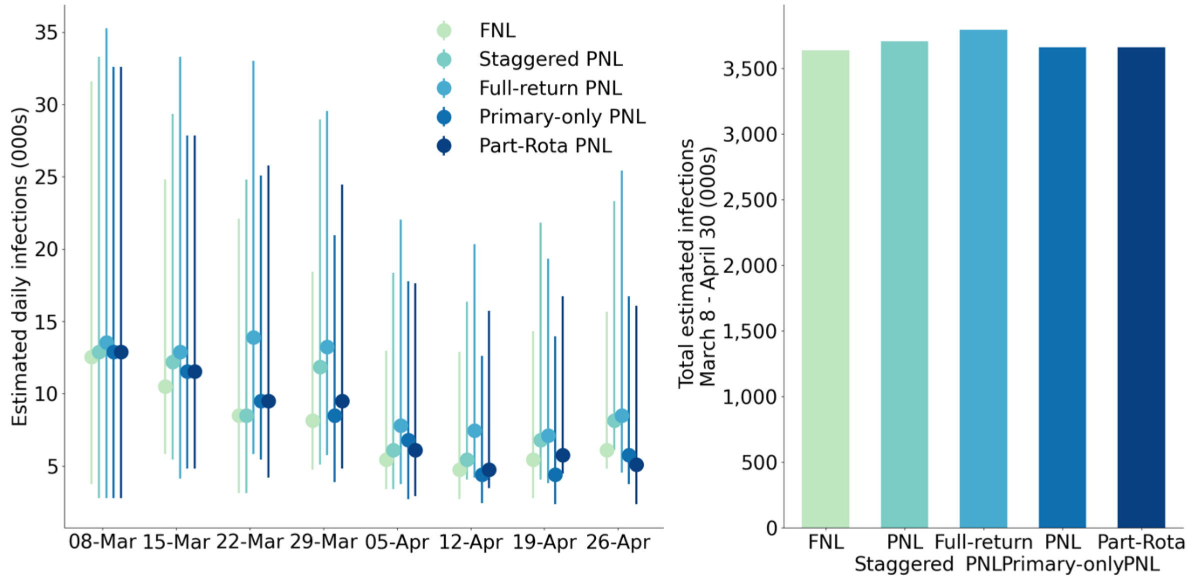


**Figure S6:** Model predicted estimated daily (a) and total (b) infections in the period from March 8 to April 30, 2021 from the calibrated model under the assumption that susceptibility of all ages is the same and that community transmission remains the same as in November lockdown between March 8,2021 and April 19, 2021 and across the five different scenarios: FNL, Staggered PNL, Full-return PNL, Primary-only PNL and Part-Rota PNL. In (a) we show the point estimates as well as the uncertainty range around this for each scenario.

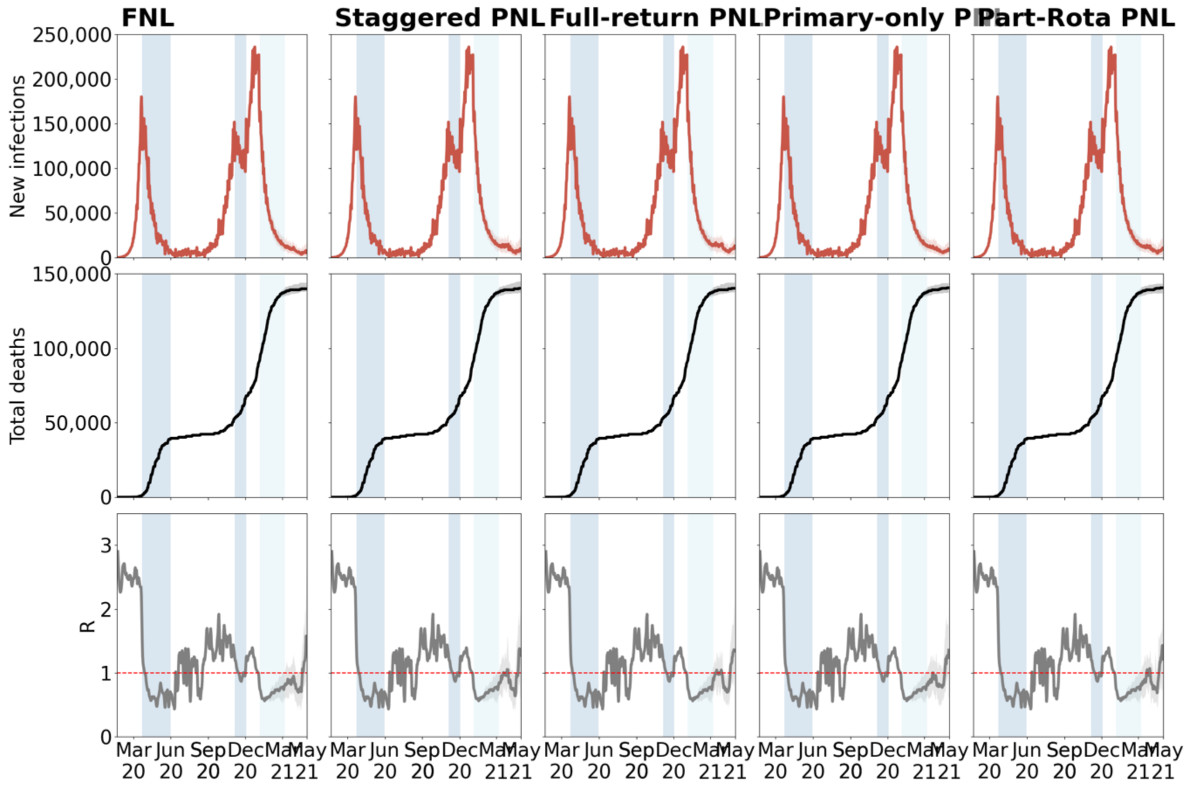


**Figure S7:** Model-predicted epidemic trajectories of the calibrated model until April 30, 2021 across FNL and PNL with different reopening strategies and under the assumption that susceptibility of all ages is the same but community transmission is 10% higher than during November lockdown between March 8,2021 and April 19, 2021. Medians across 30 simulations are indicated by solid lines and the 25% and 75% quantiles by shading. The blue bands represent the past and current national lockdowns.

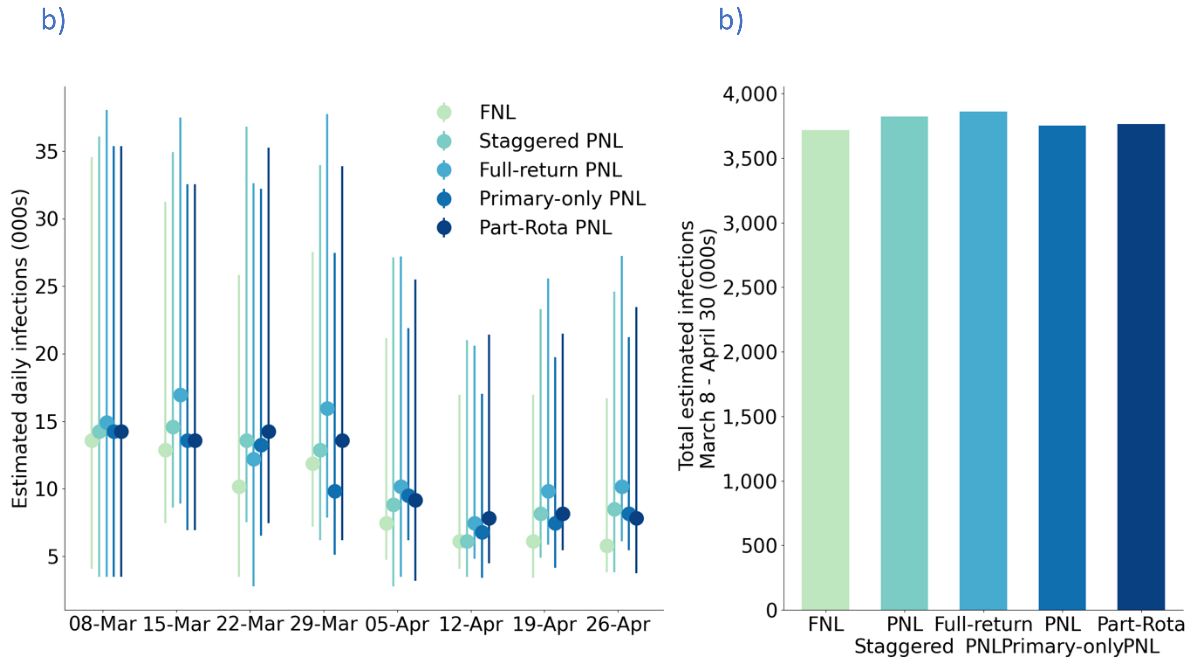


**Figure S8:** Model predicted estimated daily (a) and total (b) infections in the period from March 8 to April 30, 2021 from the calibrated model under the assumption that susceptibility of all ages is the same and that community transmission but community transmission is 10% higher than during November lockdown between March 8,2021 and April 19, 2021 and across the five different scenarios: FNL, Staggered PNL, Full-return PNL, Primary-only PNL and Part-Rota PNL. In (a) we show the point estimates as well as the uncertainty range around this for each scenario.
